# Large Language Models Readability Classification: A Variability Analysis of Sources and Metrics

**DOI:** 10.64898/2026.02.20.26346638

**Authors:** Hector Gabriel Corrale de Matos, Jan-Willem A. Wasmann, Thais Catalani Morata, Kátia de Freitas Alvarenga, Lilian Cássia Bórnia Jacob

## Abstract

Accurate health information is ineffective if patients cannot understand it. Large Language Model (LLM) health research values veridical precision; however, linguistic accessibility remains an under-examined component of output quality and usability. This study investigated two sources of variability in readability classification: differences across LLM systems and across readability metrics. The analysis tested 1,120 data points from seven systems in English and Portuguese, comparing baseline responses with a Wikipedia-grounded condition. Content was assessed using five standard readability metrics that measure distinct aspects of text complexity. Systems were statistically homogeneous at baseline but became significantly heterogeneous under Wikipedia grounding, indicating variability in the combination of Retrieval-Augmented Generation (differential readability effects of the same source-grounding instruction across systems). Significant metric variability was observed in all conditions, showing that readability metrics are not interchangeable. Although retrieval grounding is commonly used to improve accuracy, our findings show a trade-off: verified-source grounding can yield inconsistent readability. Therefore, evaluation protocols should use transparent, vendor-agnostic criteria, with metric-specific and language-aware thresholds, and be applied whenever models or grounding configurations change to support accessible cross-language health communication.

## Introduction

The application of generative artificial intelligence into medicine and audiology is an emerging trend across research and clinical settings (Lesica et al., 2021; Park et al., 2024; Peng et al., 2023; Younis et al., 2024). Scholars also contend that the prevailing momentum in artificial-intelligence research is driven less by clinical demand than by a corporate-driven narrative of inevitability, whose large-scale large language models (LLMs) extraction threatens collaborative knowledge platforms such as Wikipedia (Monett & Grigorescu, 2024; Vetter et al., 2025). Chatbots such as ChatGPT have emerged as promising tools to improve the identification, treatment, and prevention of hearing loss by providing accessible information to both patients and hearing health professionals (Aggarwal et al., 2023; Clusmann et al., 2023; Crowson, 2020; Swanepoel et al., 2023; Swanepoel & Manchaiah, 2024; Wasmann et al., 2022; Wilson et al., 2022). However, the accuracy of chatbot-generated content is still questionable, particularly the absence of thorough readability assessments (Milne-Ives et al., 2020; Sallam, 2023; Seitz et al., 2022; Shool et al., 2025).

By 2050, an estimated 2.5 billion individuals worldwide are projected to experience hearing loss (Wilson et al., 2022; World Health Organization, 2021). This trend is expected to result in significant economic and social burdens, particularly in regions with insufficient healthcare infrastructure (Jiang et al., 2023; McDaid et al., 2021). At present, approximately 430 million people require ear and hearing care globally; however, between 77% and 90% of these individuals do not have adequate access to rehabilitation services, primarily due to limited resources and insufficient information (Orji et al., 2020; World Health Organization, 2021). The lack of accessible information substantially delays timely interventions, thereby intensifying the global impact of hearing loss (Tran et al., 2021; Tsimpida et al., 2024; Wilson et al., 2017). Therefore, the development and implementation of effective, evidence-based technologies and methodologies for disseminating hearing health information are critical to addressing these challenges (Ratanjee-Vanmali et al., 2019; World Health Organization, 2021).

LLMs are deep neural network architectures utilizing the Transformer framework, trained on extensive text corpora to model and generate human-like language (Brown et al., 2020; Vaswani et al., 2017). Models such as ChatGPT, developed by OpenAI, are designed to interpret and generate human language based on comprehensive training from diverse textual sources, including websites, technical literature, and datasets (Amaratunga, 2023; Liu et al., 2023; Paaß & Giesselbach, 2023; Zhao et al., 2023). Despite significant advancements demonstrated by LLMs such as Anthropic’s Claude, Google’s Gemini, and DeepSeek, persistent accuracy issues remain. While LLMs often perform well in structured technical domains such as programming, their outputs in health-related contexts may still contain critical inaccuracies, posing risks to patient safety (Ayers et al., 2023; Singhal et al., 2023). Misinformation or fabricated content generated by these systems is commonly referred to as “artificial intelligence hallucinations” (Ji et al., 2023). In health applications, incorrect outputs can compromise medical safety by providing inappropriate recommendations (Baber et al., 2024; Hatem et al., 2023; Stojanov, 2023; Thirunavukarasu et al., 2024; Yan et al., 2023). Techniques such as retrieval-augmented generation (RAG), which incorporate contextually relevant and verified data sources, have been proposed to mitigate inaccuracies and enhance reliability, particularly in biomedical contexts (Luo et al., 2024; Shi et al., 2024; Soong et al., 2024). The rapid implementation of LLMs without addressing intrinsic limitations necessitates an urgent shift in strategy to ensure the critical evaluation of LLM-generated content uses (Monett & Grigorescu, 2024; Monett & Paquet, 2025).

Crowdsourced and open knowledge repositories, such as Wikipedia, and peer-reviewed databases, including Scopus, have been incorporated into retrieval-augmented generation (RAG) frameworks to improve the quality of information produced by LLMs (Aguilera Cora et al., 2024; Lewis et al., 2020; Liesenfeld et al., 2023; Siriwardhana et al., 2023). Wikipedia plays a significant role in reducing global knowledge disparities by providing access to scientifically validated health information (Ayers et al., 2023; Heilman et al., 2011; Kane & Ransbotham, 2016; World Health Organization, 2020). Furthermore, the development of non-Western language models and initiatives originating from the Global South has received increasing attention, highlighting ongoing efforts to expand linguistic diversity and representation in LLM technologies, exemplified by Maritaca-10B (MaritacaAI, 2026).

Although addressing “hallucinations” is essential for ensuring factual accuracy, reliability in health communication comprises both accuracy and linguistic accessibility. Information must be both correct and understandable. When patients are unable to comprehend accurate guidance, health literacy gaps may widen, and interventions may be delayed. Even highly readable false information creates considerable risks. This study shifts focus from the extensively studied issue of accuracy to the less explored aspect of LLMs’ readability, with particular emphasis on the automated assessment of generated text complexity.

This study evaluates the readability classifications of hearing health information generated by seven LLMs in both Portuguese and English. The findings aim to inform strategies for enhancing cross-language consistency in health communication. Analyses are organized into four groups based on language (Portuguese or English) and generation setting (baseline generation or Wikipedia-sourced generation). Two dimensions of homogeneity are examined: (H1) variation in classification outcomes across LLMs (model families or providers) and (H2) variation across readability metrics. The study investigates whether the choice of LLM, generation setting, or readability metric introduces systematic variability in the classification of identical health content.

## Methods

This cross-sectional study assessed the readability of text generated by seven LLM-based chat systems in response to structured audiology-related prompts in Portuguese and English. The study design and reporting followed the CHART (Chatbot Health Advice studies) guideline to ensure methodological transparency and reproducibility. Models were selected (available as of April 2025) based on two criteria: (i) they were accessible in both languages through public web interfaces that were either free to use or usage-limited (e.g., quota-restricted access, rate limits, or capped daily messages), and (ii) they represented a heterogeneous set of LLM systems from different developers and regions.

This study is exempt from formal ethical approval under the Brazilian National Health Council Resolution No. 510 of April 7, 2016, since it relied solely on the analysis of artificially generated texts, without the participation of human subjects.

The following seven LLMs were evaluated: OpenAI-ChatGPT-4o (GPT), DeepSeek-R1 (DpS), Claude-3.7-Sonnet (Cd), Google-Gemini-2.0-Flash (Gm), Mistral-AI-LeChat-8×22B (leC), Maritaca-10B-Sábia3 (Ma), and Microsoft-Copilot-April-2025 (Co).

The study employed seven LLMs and five readability metrics to investigate two dimensions of homogeneity (H1 and H2). This multi-vector approach was essential to determine whether variability in readability classifications originated from inherent differences among model architectures (H1) or from disparities among the measurement tools themselves (H2). This design thus allows for the distinction between variations in content quality (model properties) and variations in classification (metric properties).

The prompts were developed based on the World Health Organization (WHO) document titled “Hearing Aid Service Delivery Approaches for Low-and Middle-Income Settings” which served as reference material for prompt development (World Health Organization, 2024). This involved: (i) empowerment, through thematic analysis of WHO recommendations for hearing aid delivery for adults (≥18 years) and children (>5 years), which focuses on training non-specialist personnel in resource-limited settings; and (ii) appropriate referrals, by the identification of core clinical referral criteria necessitating specialist intervention during assessment or follow-up. The general recommendations provided in the WHO document were systematically converted into specific, structured prompts designed to elicit detailed clinical guidance from the LLMs.

Prompts in English (EN) and Portuguese (PT) were used to assess LLMs across eight clinical domains in hearing health, based on the WHO document. Initially composed in English, prompts were translated and culturally adapted into Portuguese by bilingual audiology specialists to ensure contextual similarity. Responses were generated exclusively from each model’s intrinsic pre-trained knowledge, without any external information retrieval, to establish a true baseline of their generative capabilities. To avoid carry-over effects (i.e., data leakage), each prompt was submitted in a separate session to ensure that each evaluation was fully independent. Additionally, the shared learning configuration across interactions was disabled because the data-sharing option intended to improve the model was disabled in the LLM used.

A thematic analysis then classified the referral criteria into eight core clinical domains: congenital anomalies, ear trauma, chronic otitis media, tympanic membrane perforation, progressive hearing loss, unilateral hearing loss, tinnitus, and severe hearing loss. In a subsequent phase, models were instructed to integrate information from Wikipedia. This orientation was made by prompt instruction. It sourced the information from the Wikipedia website in both language versions. Supplementary file 1 lists the utilized prompts.

The study generated and analyzed a corpus of 224 unique texts (112 in English and 112 in Portuguese), derived from 7 LLMs responding to 8 prompts across 2 conditions (Baseline and Wikipedia-sourced). As each text was processed by 5 readability metrics, the results yielded a total dataset of 1120 computational data points (N=280 per group) for the subsequent homogeneity analysis. Supplementary file 3 compiles the raw groups’ computational data points.

The five metrics selected for the homogeneity analysis were: i) Flesch Reading Ease (FRE): scores range from 0 to 100; higher scores indicate greater ease of reading; ii) Flesch-Kincaid Grade Level (FKG): estimates the education level required to comprehend the text; iii) Simple Measure of Gobbledygook (SMOG): focuses on polysyllabic words as indicators of complexity; iv) Automated Readability Index (ARI): based on characters per word and words per sentence; and v) Coleman-Liau Index (CLI): uses character and sentence counts to determine grade level.

This specific set of metrics was picked to ensure a strong, multidimensional assessment suitable for health-related content in a multilingual context. FRE and SMOG are widely recognized as standards for evaluating patient education materials, as they directly address the complexity of medical terminology (Singh et al., 2024). To reduce linguistic biases inherent to syllable-based formulas when comparing English and Portuguese, ARI and CLI were included; these metrics rely on character counts, providing a more stable baseline for cross-language variability analysis, as endorsed by recent literature (Okuhara et al., 2025).

Readability calculations and subsequent descriptive and inferential statistical analyses were performed in Python (version 3.12.0) within the JupyterLab environment (version 4.5.2). Readability metrics were computed using specific libraries: alt-legibilidade (version 0.1.2; ALT, 2025) for Portuguese texts, following the ATL framework (Moreno et al., 2022), and textstat (version 0.7.12; Bansal & Aggarwal, 2025) for English texts. The reference scales for each metric in both languages followed the definitions of the respective libraries. The SMOG index in Portuguese was calculated using a custom function based on the regression formula (McLaughlin, 1969), adapted for Brazilian Portuguese syllable counting using the pyphen library (version 0.17.2; Ayoub, 2025), due to the lack of native library support for the SMOG formula in Portuguese. Prompt responses for each LLM, in both English and Portuguese, were analyzed using the readability metric formula for each language. The text responses and derived data points of the readability scores were recorded in a spreadsheet using the LibreOffice Calc application (version 25.8). The prompts were generated and the data collected between April 25 and 27, 2025, in São Paulo, Brazil. Supplementary file 2 records the response texts of the LLMs in both languages in the baseline and Wikipedia-oriented schemas.

While readability metrics applied to the same corpus are theoretically correlated, we treated each classification instance as an independent observation for the homogeneity analysis. This approach was chosen because (1) the metrics utilize distinct algorithmic inputs (characters vs. syllables vs. lexicons), creating operational independence; and (2) aggregating these scores (e.g., using majority voting or averaging) was explicitly avoided because it would necessitate treating inter-metric disagreement as statistical noise to be eliminated. Aggregation would artificially homogenize the data, obscuring the metric variability that this study aims to reveal and quantifying false consensus where none exists.

The G-test (likelihood-ratio test) was utilized to evaluate the study’s hypotheses regarding the homogeneity of classifications (statistical significance at p <.05). This test is designed to compare proportions of nominal categories across different groups. The use of this test is predicated on the independence of observations, a condition met by our data as each classified item represents a distinct entity and is counted in only one cell. The binarization thresholds were selected to distinguish between general public communication and specialized academic discourse, rather than establishing an optimal health literacy target. This statistical approach necessitated the binarization of the readability metric outputs into two nominal categories:’Upper’ (representing lower complexity and higher readability) and’Lower’ (representing higher complexity and lower readability). The binarization process in the nominal categories was performed using the LibreOffice Calc application (version 25.8). Readability scores indicating lower complexity (FRE ≥ 30 or FKGL/SMOG/ARI/CLI ≤ 12) were classified as “Upper,” while scores indicating higher complexity (FRE < 30 or FKGL/SMOG/ARI/CLI > 12) were classified as “Lower.” According to the Flesch scale classifications, scores between 0–30 represent’Very Difficult’ texts typical of scientific journals; therefore, the cut-off of 30 serves as a boundary for excluding highly technical characteristics (Doak et al., 1985; Flesch, 1948). Similarly, patient education materials target 6th–8th grade levels as suggested; 12th-grade threshold aligns with the completion of secondary education (Kutner et al., 2006). The analysis was then stratified into four independent groups: Portuguese-Baseline, Portuguese-Wikipedia, English-Baseline, and English-Wikipedia. To test the two main hypotheses, two distinct analytical approaches were applied to the generated contingency tables. For the H1 test (Model Architecture Influence), data was structured by LLM, creating four 2×7 contingency tables (total N=280 per group). This design tested whether the response proportions were homogeneous across the seven models, with each model contributing 40 data points derived from 8 prompts and 5 metrics. For the H2 test (Metric Homogeneity), data was grouped by the readability metric to determine if the tools classified content consistently. This resulted in four 2×5 contingency tables (total N=280 per group). This design was tested for homogeneous response proportions across the five metrics, with each metric contributing 56 data points (7 models x 8 prompts). For each test, Cramér’s V (V ≈ 0.1: small; V ≈ 0.3: medium; V ≈ 0.5: large) was calculated as a measure of effect size (strength of association), and 95% Confidence Intervals (CI) were calculated for the proportion of “Upper” responses to assess the precision of the estimates. For any test group found to be statistically significant (p <.05), a post-hoc analysis of Standardized Residuals (SR) was performed, using |SR| > 1.96 to identify the specific cells (models or metrics) that significantly contributed to the overall difference. Additionally, 95% binomial confidence intervals (Clopper-Pearson exact method) were calculated to assess the precision of the observed proportions for each response level. Statistical analysis was conducted in R (version 4.4.2) on the RStudio platform. Supplementary file 3 documents all categorization tables, frequency counts of category occurrences, and contingency tables by model (Baseline and Wikipedia-sourced) and language (Portuguese and English).

All code, datasets, statistical analysis, and research supplementary materials were deposited in the Open Science Framework repository (10.17605/OSF.IO/QCXPS), adhering to the FAIR (Findable, Accessible, Interoperable, and Reusable) data principles established by the Research Data Alliance and the Committee on Data of the International Science Council.

## Results

To test our first hypothesis, H1 (Model Architecture Influence), we first analyzed whether readability classifications were homogeneous across the seven LLMs. Table 1 details these findings. Significant variability among models was observed in the two Wikipedia-sourced groups (Portuguese-Wikipedia: p=.0027; English-Wikipedia: p=.0011), but not in the baseline groups (Portuguese-Baseline: p=.1134; English-Baseline: p=.2255). Post-hoc analysis (Standardized Residual) of the significant groups identified the specific models that contributed to this variance.

**Table 1.** G-test for homogeneity for response levels to readability scores (“Lower” vs. “Upper”) across LLMs, stratified by language (Portuguese and English) and source (Baseline and Wikipedia-sourced). Each cell displays the proportion (%), raw frequency (n), 95% Confidence Interval [CI], and Standardized Residual (SR). Overall group statistics include the G-test value (G), p-value, and Effect Size (Cramér’s V). SR values are shown only for significant tests (p < 0.05); Not Applicable (NA) is used for non-significant groups.

| G | p-value | V | Level | GPT | DpS | Cd | Gm | leC | Ma | Co |
| --- | --- | --- | --- | --- | --- | --- | --- | --- | --- | --- |
| Portuguese - Baseline |  |  |  |  |  |  |  |  |  |  |
| 10.278 | 0.1134 | 0.1790<br>[0.13-0.31] | Lower | 77.5% (31)<br>[62.5-87.7]<br>SR: NA | 87.5% (35)<br>[73.9-94.5]<br>SR: NA | 72.5% (29)<br>[57.2-83.9]<br>SR: NA | 77.5% (31)<br>[62.5-87.7]<br>SR: NA | 82.5% (33)<br>[68.1-91.3]<br>SR: NA | 95% (38)<br>[83.5-98.6]<br>SR: NA | 80% (32)<br>[65.2-89.5]<br>SR: NA |
|  |  |  | Upper | 22.5% (9) SR:<br>NA | 12.5% (5) SR:<br>NA | 27.5% (11)<br>SR: NA | 22.5% (9)<br>SR: NA | 17.5% (7)<br>SR: NA | 5% (2)<br>SR: NA | 20% (8)<br>SR: NA |
| Portuguese - Wikipedia-sourced |  |  |  |  |  |  |  |  |  |  |
| 20.023 | 0.0027 | 0.2503<br>[0.18-0.38] | Lower | 85% (34)<br>[70.9-92.9]<br>SR: 0.28 | 72.5% (29)<br>[57.2-83.9]<br>SR: -0.60 | 72.5% (29)<br>[57.2-83.9]<br>SR: -0.60 | 67.5% (27)<br>[52.0-79.9]<br>SR: -0.95 | 87.5% (35)<br>[73.9-94.5]<br>SR: 0.45 | 85% (34)<br>[70.9-92.9]<br>SR: 0.28 | 97.5% (39)<br>[87.1-99.6]<br>SR: 1.15 |
|  |  |  | Upper | 15% (6)<br>SR: -0.57 | 27.5% (11)<br>SR: 1.25 | 27.5% (11)<br>SR: 1.25 | 32.5% (13)<br>SR: 1.97 | 12.5% (5)<br>SR: -0.93 | 15% (6)<br>SR: -0.57 | 2.5% (1)<br>SR: -2.39 |
| English - Baseline |  |  |  |  |  |  |  |  |  |  |
| 8.175 | 0.2255 | 0.1699<br>[0.11-0.32] | Lower | 62.5% (25)<br>[47.0-75.8]<br>SR: NA | 80% (32)<br>[65.2-89.5]<br>SR: NA | 87.5% (35)<br>[73.9-94.5]<br>SR: NA | 77.5% (31)<br>[62.5-87.7]<br>SR: NA | 75% (30)<br>[59.8-85.8]<br>SR: NA | 70% (28)<br>[54.6-81.9]<br>SR: NA | 77.5% (31)<br>[62.5-87.7]<br>SR: NA |
|  |  |  | Upper | 37.5% (15)<br>SR: NA | 20% (8)<br>SR: NA | 12.5% (5)<br>SR: NA | 22.5% (9)<br>SR: NA | 25% (10)<br>SR: NA | 30% (12)<br>SR: NA | 22.5% (9)<br>SR: NA |
| English - Wikipedia-sourced |  |  |  |  |  |  |  |  |  |  |
| 22.178 | 0.0011 | 0.2866<br>[0.19-0.43] | Lower | 50% (20)<br>[35.2-64.8]<br>SR: -1.91 | 80% (32)<br>[65.2-89.5]<br>SR: 0.26 | 92.5% (37)<br>[80.1-97.4]<br>SR: 1.16 | 72.5% (29)<br>[57.2-83.9]<br>SR: -0.28 | 77.5% (31)<br>[62.5-87.7]<br>SR: 0.08 | 82.5% (33)<br>[68.1-91.3]<br>SR: 0.44 | 80% (32)<br>[65.2-89.5]<br>SR: 0.26 |
|  |  |  | Upper | 50% (20)<br>SR: 3.44 | 20% (8)<br>SR: -0.47 | 7.5% (3)<br>SR: -2.09 | 27.5% (11)<br>SR: 0.51 | 22.5% (9)<br>SR: -0.14 | 17.5% (7)<br>SR: -0.79 | 20% (8)<br>SR: -0.47 |

**Note:** Upper= lower complexity and higher readability, Lower=higher complexity and lower readability. OpenAI-ChatGPT-4o (GPT), DeepSeek-R1 (DpS), Claude-3.7-Sonnet (Cd), Google-Gemini-2.0-Flash (Gm), Mistral-AI-LeChat-8x22B (leC), Maritaca-10B-Sábia3 (Ma), and Microsoft-Copilot-April-2025 (Co). **Source:** Created by the authors.

To address H2 (Metric Homogeneity), we tested whether the five readability scores were homogeneous in their classifications (see Table 2). In a finding that mirrors the architectural analysis, significant variability and medium effect sizes (V = 0.22 to 0.37) were observed among metrics in all four groups. This indicates that the choice of readability metric introduces a significant source of classification variability, independent of the model or language.

**Table 2.**
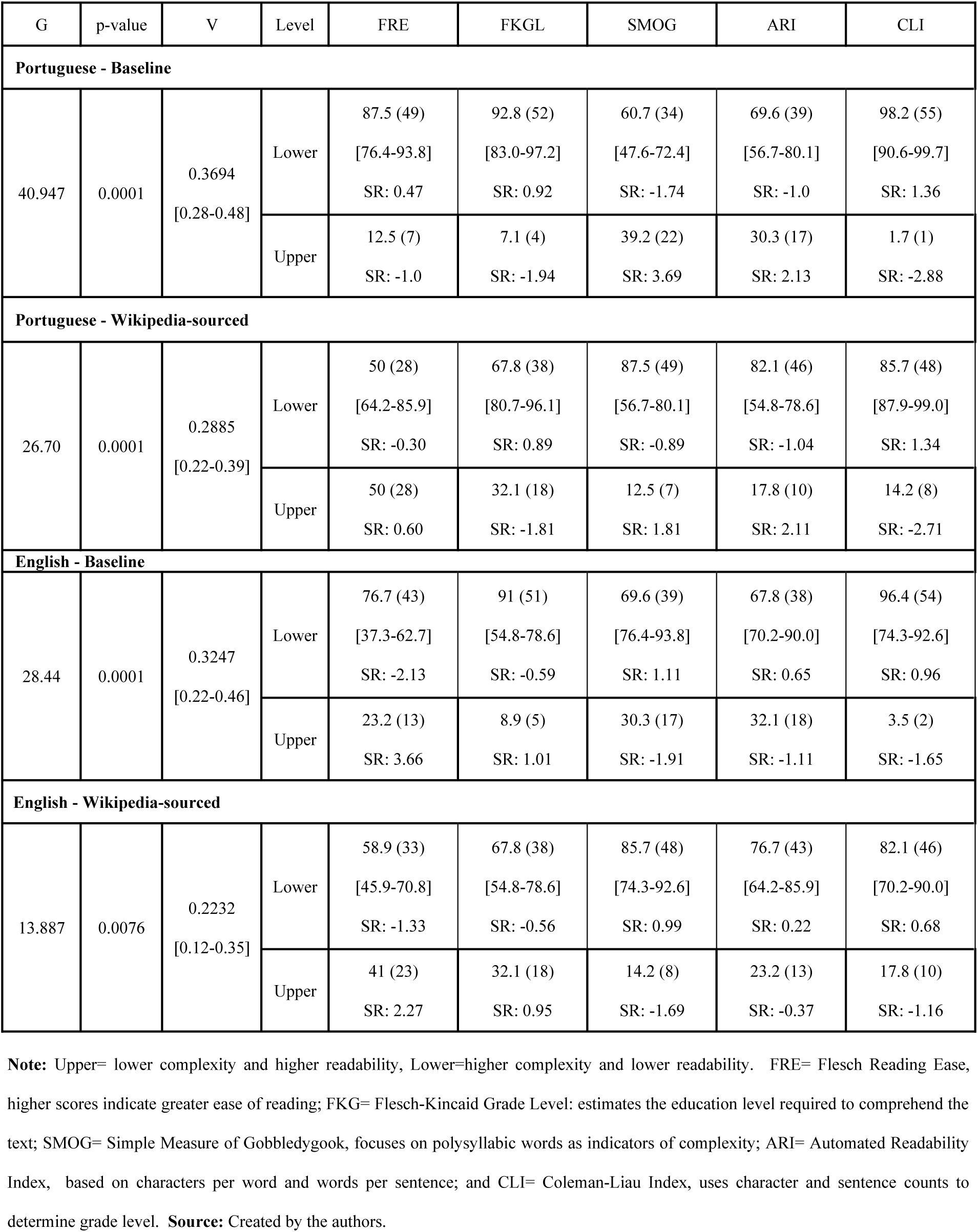
G-test for homogeneity comparing the proportion of “Lower” vs. “Upper” responses for readability scores across five metrics (FRE, FKGL, SMOG, ARI, and CLI), stratified by language (Portuguese and English) and source (Baseline and Wikipedia-sourced). Each cell displays the response proportion (%), raw frequency (n), and 95% Confidence Interval [CI]. The overall test statistics (G-statistic, p-value, and Effect Size/Cramér’s V) and Post-hoc test (Standardized Residual) are shown for each group.

## Discussion

This study investigated two dimensions of variability in readability classifications of LLM-generated hearing health information: model design effects (H1) and metric consistency (H2). The data confirm that readability variability results from multiple sources: significant heterogeneity was found both among model designs and among the readability metrics themselves.

### Model Architecture Variability

Analysis shows that between-model differences in readability classifications depend on the generation setting (H1). Under baseline conditions (generation from pre-trained knowledge only), readability classifications were statistically homogeneous across the seven LLM systems in both Portuguese and English (Table 1). This pattern suggests that, in a standard chat-style generation setting, instruction-following and alignment objectives that target clear, beneficial responses may yield broadly similar surface-level readability profiles across systems, even free of explicit optimization for readability formulas.

These findings show that identical knowledge augmentation produces heterogeneous readability outcomes. This phenomenon is defined in this study as’integration variability’: the differential processing of identical external knowledge sources by different model designs. The baseline homogeneity indicates that models may be comparably tuned for general conversational output. The subsequent emergence of heterogeneity in the Wikipedia-sourced condition is relevant, given that these models are trained on Wikipedia data. It is hypothesized that this shift remains a function of the retrieval-augmented generation (RAG) implementation itself. In baseline, models may optimize for conversational simplicity. However, the Wikipedia-sourced condition forces the models to retrieve, prioritize, and synthesize the provided source content. If this source corpus is, on average, more linguistically complex than the model’s default generative output, the forced integration would predictably increase overall complexity. The noted variability would then arise because different architectures employ distinct internal mechanisms to execute this forced synthesis; some may tend to summarize (reducing complexity), while others may rewrite or quote more directly (preserving the source’s complexity).

Wikipedia-sourcing was not a uniform intervention in our prompt-based implementation. Even under identical instructions to ground responses in Wikipedia, LLMs can differ in how they integrate the provided source, e.g., the degree of paraphrasing, summarization, quotation, and (in Portuguese) adaptation/translation, which can amplify surface-level differences captured by formula-based readability metrics. This helps explain why grounding on a verified source can maintain, or even increase, between-model variability in readability outcomes, notably under binary classification thresholds. In practice, the results point to a governance trade-off: strengthening verifiability through source grounding may unintentionally weaken accessibility. This illustrates the risks of’techno-solutionism,’ the assumption that integrating AI tools automatically resolves complex problems without causing new harms (Monett & Paquet, 2025). Therefore, post-deployment protocols must jointly audit factual reliability and readability.

Homogeneity disappeared when models were augmented with Wikipedia-sourced knowledge via prompt-based source grounding. Considerable variability emerged in both Portuguese-Wikipedia and English-Wikipedia conditions (Table 1). Post-hoc standardized residual analysis identified the specific models driving this variation. In the English-Wikipedia group, GPT produced significantly more low-complexity content (50% classified as Upper), while Claude generated more complex content (92.5% classified as Lower). Similarly, on Portuguese Wikipedia, Copilot produced markedly less comprehensible content (97.5% in the Lower category), whereas Gemini trended toward greater accessibility (32.5% in the Upper category). These data demonstrate that a standardized Wikipedia-grounding instruction can produce heterogeneous readability outcomes, consistent with integration variability across LLM systems.

This cross-language shift in the models driving variability suggests an interaction between language and source-grounding: the readability impact of Wikipedia-sourcing is not uniform across LLM systems that may depend on (i) how each system integrates the source under the prompt-based grounding procedure and (ii) language-specific properties of the generated output and the underlying Wikipedia edition. These mechanisms are plausible but were not directly measured in this study and warrant controlled follow-up analyses.

### Metric Variability

Rejecting our second hypothesis (H2), the five readability metrics showed heterogeneity in their categorizations across all conditions (Baseline and Wikipedia-sourced). Post-hoc analysis (see Table 2) shows that such variation is driven by computational differences; for example, SMOG (syllable-based) metrics frequently classify content differently than CLI (character-based) metrics. The result contests the assumption that these metrics are interchangeable tools for assessing LLMs’ output. The findings show empirically that this interchangeable use is methodologically unsound for evaluating LLM outputs. The choice of how to measure readability creates a source of variability as significant as the choice of which LLMs to use. The data assesses consistency, not accuracy. Without checking against human insight, no single metric can be identified as superior or more meaningful. Therefore, upcoming research should use a battery of metrics rather than relying on a single score to avoid systematic bias.

### Comparison with Prior Research and Implications

The findings broaden the current literature by identifying linguistic accessibility as an essential factor in LLM accuracy. Previous studies have documented differences in LLM-generated medical content, primarily concentrating on factual accuracy and “hallucinations” (Baber et al., 2024; Thirunavukarasu et al., 2024), as well as on the degradation of health information integrity in the current infodemic (Eysenbach, 2020; World Health Organization, 2020). The research uncovers a parallel dimension of variability: linguistic accessibility. Although RAG is used to improve accuracy (Luo et al., 2024; Shi et al., 2024), our results show a trade-off: inconsistent readability. It was found that adding external knowledge makes text complexity unpredictable, with significant variation across models.

This “integration variability” has significant implications for global health equity, especially in settings with low health literacy. This challenge, analogous to “learning poverty” (The World Bank et al., 2022) in global education, can significantly inhibit access to care. The case examined by our study was the public health issue of hearing loss. Given its significant global burden (World Health Organization, 2021, 2024), with 77–90% of those requiring rehabilitation lacking adequate access to care (Orji et al., 2020), the linguistic accessibility of digital health information is a critical determinant of access to health services and interventions. The real-world impact on patients is direct: information that is too complex to understand significantly delays the time to seek care, identify hearing disorders, and initiate needed interventions. The findings suggest that choosing different LLM systems may unintentionally create new health literacy disparities. The impact can also vary by language. For example, one system may consistently produce more complex text that is harder to understand, while another may produce more accessible text. This highlights the need for clear criteria when selecting models and readability metrics to evaluate hearing health information.

Furthermore, our data show that improving readability should be addressed primarily at the provider and system levels by designing and validating LLM-generated health information that is consistently understandable across models and languages. Simultaneously, broader educational curricula should incorporate health literacy and foundational AI literacy as essential competencies to enable individuals to critically interpret LLM-mediated information. Such measures are consistent with the’critical AI literacy’ approach to enable users in handling the accounts and artificial information ecosystem procured by LLMs (Monett & Grigorescu, 2024). These approaches are complementary: making information easier to understand reduces avoidable barriers in the short term, while education may yield wider long-range benefits beyond AI-specific contexts.

The metric heterogeneity we observed (H2) supplies methodological clarity. While the literature has noted the theoretical limitations of readability formulas (Collins-Thompson, 2014), our findings show that these metrics also lack practical consistency when applied to LLM-generated content. This variation implies that researchers cannot securely deploy these tools interchangeably, as metric selection itself brings about a significant source of systematic bias into readability assessments. This finding is notably concerning for cross-linguistic research, as the variability was persistent in both Portuguese and English.

The post-hoc analysis conducted expands on concerns raised in the literature regarding LLM reliability (Milne-Ives et al., 2020; Sallam, 2023). The data show that adding a Wikipedia grounding step does not yield uniform readability effects across LLMs. Therefore, these systems should not be treated as interchangeable in practice. Instead, readability evaluation should rely on universal, model-agnostic criteria that can be applied consistently across vendors, languages, and deployment settings. Under this approach, models that fail to meet the predefined readability targets under the same evaluation protocol should be avoided (or constrained) for patient-facing health communication. In other words, the implication is not that each model requires a different tool, but that a single solid evaluation framework is needed to screen and monitor all candidate systems, especially when grounding is introduced.

This design specificity underscores the necessity of structured knowledge production on crowdsourced platforms. Wikipedia, an open and collaboratively curated health information source, was officially recognized as a Digital Public Good by the Digital Public Goods Alliance in 2025. It now serves as a critical foundation for source-grounded (RAG-like) systems (Vetter et al., 2025; Wikimedia Foundation, 2025). This assimilation brings notable epistemic risks. Recent scholarship warns of a’circular dependency’ or’Ouroboros effect,’ in which LLMs trained on generated content, growing prevalent on Wikipedia, may suffer from’model collapse’ and reduced epistemic diversity (Shumailov et al., 2024; Wright et al., 2025). Furthermore, the uncritical automated ingestion of this corpus renders RAG systems vulnerable to’data poisoning’ and vandalism, possibly amplifying misinformation at scale without the safeguards of human contextualization (Alber et al., 2025).

Therefore, investing in the human-centric accessibility and integrity of this core corpus constitutes a necessary public health strategy to improve and standardize the readability of LLM-generated health information. Treating resources as public goods is essential to prevent the commodification of knowledge, disrupt the recursive cycle of synthetic hallucinations, and ensure equitable access (Monett & Paquet, 2025).

### Limitations and Future Directions

The methodological scope introduces limitations due to the dichotomizing of readability scores (“Upper” vs. “Lower”), which enabled homogeneity testing but masked small differences in text complexity; continuous or ordinal analyses could capture finer between-model variation. Furthermore, our’Wikipedia-sourced’ condition was implemented by prompting the LLMs to the Wikipedia source, rather than within a formal computational RAG system. This methodological decision was deliberate, designed to isolate the models’ integration and synthesis capabilities from automated retrieval. However, this methodological-based choice acts as a key alternative explanation for our H1 results. The observed’integration variability’ may originate not only from the models’ internal processing but from the specific prompt-based method used to instruct them to integrate information from the source. Subsequent research should validate these findings using dedicated RAG frameworks employed to determine whether this variation persists when retrieval is automated.

Additionally, the context on hearing health prompts caution when extrapolating to other medical specialties.

While this narrow focus ensures high clinical relevance and content validity (as the prompts were derived from WHO guidelines), the observed variability in integration may be domain-dependent and warrant validation across multiple clinical contexts.

This data represents a temporal snapshot of seven specific LLM architectures; given the swift pace of model iteration, longitudinal research is needed to assess the steadiness of these readability profiles. Moreover, our results are specific to the Wikipedia-sourced implementation; alternative knowledge sources (e.g., peer-reviewed databases) or retrieval configurations may yield different heterogeneity patterns.

The readability metrics employed assess linguistic complexity instead of clinical validity or semantic accuracy. A critical next step is to validate these outcomes by testing them in real-world conditions with patients and clinicians. A text classified as “Upper” (easy to read) could still contain misinformation; veracity assessment evaluation, though essential, was beyond this study’s scope. The analysis was confined to English and Portuguese; architectural behaviors during knowledge integration may differ markedly in languages with different morphological structures or diminished resource representation in training data. Furthermore, this study focused on classic, formula-based readability metrics; subsequent studies ought to investigate whether these outcomes persist when using modern assessment methods, such as metrics based on transformer models or psycholinguistic principles.

It is also relevant to consider the basic structural dissonance between the knowledge source and the generative models. Wikipedia content is human-curated, reflecting structured editorial choices and human cognitive patterns. In contrast, LLMs generate text based on probabilistic token prediction. This tension demonstrates a wider’algorithmification’ (Monett & Grigorescu, 2024), in which complex human-curated narratives are reduced to algorithmic outcomes that may overlook the social value of health communication. The observed heterogeneity may, therefore, stem from this intrinsic’structural friction’: different architectures may employ different mechanisms to reconcile their probabilistic logic with the rigid, human-defined structure of the base text, a variability factor potentially independent from the specific semantic content.

## Conclusion

This study shows that variability in readability classification of LLM-generated health information has two independent sources: integration variability (H1) and metric variability (H2). Across both languages, systems were broadly homogeneous in baseline generation, yet became significantly heterogeneous under Wikipedia grounding, indicating that the same verified-source instruction can yield different readability outcomes depending on the model. In parallel, all five readability metrics disagreed systematically across conditions, confirming that metrics are not interchangeable and that the chosen metric set can materially change conclusions. Together, these outcomes make a concrete governance trade-off explicit: source grounding may improve verifiability, but it can also reduce readability and/or increase inconsistency in readability across systems. Therefore, stakeholders responsible for quality, researchers, clinicians, developers/system owners, and policy or regulatory actors should embrace transparent, provider-neutral evaluation criteria while applying metric-specific and language-aware thresholds, and repeat these audits whenever models or grounding configurations change, to avoid unintentionally amplifying cross-language health inequities.

## Statements and Declarations

### Ethics Approval

Ethics approval was not required for this study as it involved the computational analysis of data generated by artificial intelligence systems and did not involve human participants or animal subjects.

### Data Availability

The datasets generated and analysed during the current study, along with the R computational analysis and supplementary files, are available in the Open Science Framework repository (10.17605/OSF.IO/QCXPS).

## Acknowledgments

This work was supported by the Fundação de Amparo à Pesquisa do Estado de São Paulo (FAPESP) under Grant No. #2024/05572-7.

## Author Contributions. Conceptualization

HGCM, LCBJ, TCM, JWAW, and KFA contributed to the conception and design of the study. Methodology and Data Curation: HGCM developed the methodology and performed the data collection from Large Language Models. Formal Analysis: HGCM performed the statistical and computational analysis. Writing – Original Draft: HGCM. Writing – Review & Editing: HGCM, LCBJ, TCM, JWAW, and KFA read and approved the final manuscript.

## References

Aggarwal, A., Tam, C. C., Wu, D., Li, X., & Qiao, S. (2023). Artificial Intelligence–Based Chatbots for Promoting Health Behavioral Changes: Systematic Review. Journal of Medical Internet Research, 25, e40789. 10.2196/40789

Aguilera Cora, E., Lopezosa, C., & Codina, L. (2024). Scopus AI Beta: Functional analysis and cases. http://hdl.handle.net/10230/58658

Alber, D. A., Yang, Z., Alyakin, A., Yang, E., Rai, S., Valliani, A. A., Zhang, J., Rosenbaum, G. R., Amend-Thomas, A. K., Kurland, D. B., Kremer, C. M., Eremiev, A., Negash, B., Wiggan, D. D., Nakatsuka, M. A., Sangwon, K. L., Neifert, S. N., Khan, H. A., Save, A. V.,…Oermann, E. K. (2025). Medical large language models are vulnerable to data-poisoning attacks. Nature Medicine, 31(2), 618–626. 10.1038/s41591-024-03445-1

ALT. (2025). alt-legibilidade: Ferramenta para análise de legibilidade de textos em português (Version 0.1.2) [Python; OS Independent]. https://github.com/marcopolomoreno/alt-python

Amaratunga, T. (2023). Understanding Large Language Models: Learning Their Underlying Concepts and Technologies. Apress. 10.1007/979-8-8688-0017-7

Ayers, J. W., Poliak, A., Dredze, M., Leas, E. C., Zhu, Z., Kelley, J. B., Faix, D. J., Goodman, A. M., Longhurst, C. A., Hogarth, M., & Smith, D. M. (2023). Comparing Physician and Artificial Intelligence Chatbot Responses to Patient Questions Posted to a Public Social Media Forum. JAMA Internal Medicine, 183(6), 589. 10.1001/jamainternmed.2023.1838

Ayoub, G. (2025). pyphen: Pure Python module to hyphenate text (Version 0.17.2) [Python]. https://www.courtbouillon.org/pyphen

Baber, H., Nair, K., Gupta, R., & Gurjar, K. (2024). The beginning of ChatGPT – a systematic and bibliometric review of the literature. Information and Learning Sciences, 125(7/8), 587–614. 10.1108/ILS-04-2023-0035

Bansal, S., & Aggarwal, C. (2025). textstat: Calculate statistical features from text (Version 0.7.12) [Python]. https://github.com/textstat/textstat

Brown, T. B., Mann, B., Ryder, N., Subbiah, M., Kaplan, J., Dhariwal, P., Neelakantan, A., Shyam, P., Sastry, G., Askell, A., Agarwal, S., Herbert-Voss, A., Krueger, G., Henighan, T., Child, R., Ramesh, A., Ziegler, D. M., Wu, J., Winter, C.,…Amodei, D. (2020). *Language Models are Few-Shot Learners* (Version 4). arXiv. 10.48550/ARXIV.2005.14165

Clusmann, J., Kolbinger, F. R., Muti, H. S., Carrero, Z. I., Eckardt, J.-N., Laleh, N. G., Löffler, C. M. L., Schwarzkopf, S.-C., Unger, M., Veldhuizen, G. P., Wagner, S. J., & Kather, J. N. (2023). The future landscape of large language models in medicine. Communications Medicine, 3(1), 141. 10.1038/s43856-023-00370-1

Collins-Thompson, K. (2014). Computational assessment of text readability: A survey of current and future research. ITL - International Journal of Applied Linguistics, 165(2), 97–135. 10.1075/itl.165.2.01col

Crowson, M. G. (2020). Artificial Intelligence to Support Hearing Loss Diagnostics. The Hearing Journal, 73(11), 8,9. 10.1097/01.HJ.0000722496.35603.8a

Doak, C. C., Doak, L. G., & Root, J. H. (1985). Teaching patients with low literacy skills. Lippincott.

Eysenbach, G. (2020). How to Fight an Infodemic: The Four Pillars of Infodemic Management. Journal of Medical Internet Research, 22(6), e21820. 10.2196/21820

Flesch, R. (1948). A new readability yardstick. Journal of Applied Psychology, 32(3), 221–233. 10.1037/h0057532

Hatem, R., Simmons, B., & Thornton, J. E. (2023). A Call to Address AI “Hallucinations” and How Healthcare Professionals Can Mitigate Their Risks. Cureus. 10.7759/cureus.44720

Heilman, J. M., Kemmann, E., Bonert, M., Chatterjee, A., Ragar, B., Beards, G. M., Iberri, D. J., Harvey, M., Thomas, B., Stomp, W., Martone, M. F., Lodge, D. J., Vondracek, A., De Wolff, J. F., Liber, C., Grover, S. C., Vickers, T. J., Meskó, B., & Laurent, M. R. (2011). Wikipedia: A Key Tool for Global Public Health Promotion. Journal of Medical Internet Research, 13(1), e14. 10.2196/jmir.1589

Ji, Z., Lee, N., Frieske, R., Yu, T., Su, D., Xu, Y., Ishii, E., Bang, Y. J., Madotto, A., & Fung, P. (2023). Survey of Hallucination in Natural Language Generation. ACM Computing Surveys, 55(12), 1–38. 10.1145/3571730

Jiang, C.-Y., Han, K., Yang, F., Yin, S.-Y., Zhang, L., Liang, B.-Y., Wang, T.-B., Jiang, T., Chen, Y.-R., Shi, T.-Y., Liu, Y.-C., Chen, S.-W., Tong, B.-S., Liu, Y.-H., Pan, H.-F., & Han, Y.-X. (2023). Global, regional, and national prevalence of hearing loss from 1990 to 2019: A trend and health inequality analyses based on the Global Burden of Disease Study 2019. Ageing Research Reviews, 92, 102124. 10.1016/j.arr.2023.102124

Kane, G. C., & Ransbotham, S. (2016). Content as Community Regulator: The Recursive Relationship Between Consumption and Contribution in Open Collaboration Communities. Organization Science, 27(5), 1258–1274. 10.1287/orsc.2016.1075

Kutner, M., Greenberg, E., Jin, Y., & Paulsen, C. (2006). The Health Literacy of America’s Adults: Results from the 2003 National Assessment of Adult Literacy | IES. https://nces.ed.gov/use-work/resource-library/report/statistical-analysis-report/health-literacy-americas-adults-results-2003-national-assessment-adult-literacy

Lesica, N. A., Mehta, N., Manjaly, J. G., Deng, L., Wilson, B. S., & Zeng, F.-G. (2021). Harnessing the power of artificial intelligence to transform hearing healthcare and research. Nature Machine Intelligence, 3(10), 840–849. 10.1038/s42256-021-00394-z

Lewis, P., Perez, E., Piktus, A., Petroni, F., Karpukhin, V., Goyal, N., Küttler, H., Lewis, M., Yih, W., Rocktäschel, T., Riedel, S., & Kiela, D. (2020). *Retrieval-Augmented Generation for Knowledge-Intensive NLP Tasks* (Version 4). arXiv. 10.48550/ARXIV.2005.11401

Liesenfeld, A., Lopez, A., & Dingemanse, M. (2023). Opening up ChatGPT: Tracking openness, transparency, and accountability in instruction-tuned text generators. 10.48550/ARXIV.2307.05532

Liu, Y., Han, T., Ma, S., Zhang, J., Yang, Y., Tian, J., He, H., Li, A., He, M., Liu, Z., Wu, Z., Zhao, L., Zhu, D., Li, X., Qiang, N., Shen, D., Liu, T., & Ge, B. (2023). Summary of ChatGPT-Related research and perspective towards the future of large language models. Meta-Radiology, 1(2), 100017. 10.1016/j.metrad.2023.100017

Luo, M.-J., Pang, J., Bi, S., Lai, Y., Zhao, J., Shang, Y., Cui, T., Yang, Y., Lin, Z., Zhao, L., Wu, X., Lin, D., Chen, J., & Lin, H. (2024). Development and Evaluation of a Retrieval-Augmented Large Language Model Framework for Ophthalmology. JAMA Ophthalmology, 142(9), 798. 10.1001/jamaophthalmol.2024.2513

MaritacaAI. (2026). Research. MaritacaAI. Maritaca AI Site. https://www.maritaca.ai/research

McDaid, D., Park, A.-L., & Chadha, S. (2021). Estimating the global costs of hearing loss. International Journal of Audiology, 60(3), 162–170. 10.1080/14992027.2021.1883197

McLaughlin, G. H. (1969). SMOG Grading—A New Readability Formula. J Reading, 12(8), 639–646.

Milne-Ives, M., De Cock, C., Lim, E., Shehadeh, M. H., De Pennington, N., Mole, G., Normando, E., & Meinert, E. (2020). The Effectiveness of Artificial Intelligence Conversational Agents in Health Care: Systematic Review. Journal of Medical Internet Research, 22(10), e20346. 10.2196/20346

Monett, D., & Grigorescu, B. (2024). Deconstructing the AI Myth: Fallacies and Harms of Algorithmification. European Conference on E-Learning, 23(1), 242–248. 10.34190/ecel.23.1.2759

Monett, D., & Paquet, G. (2025). Against the Commodification of Education—If harms then not AI. *Journal of Open*, Distance, and Digital Education, 2(1). 10.25619/WAZGW457

Moreno, G. C. de L., de Souza, M. P. M., Hein, N., & Hein, A. K. (2022). ALT: Um software para análise de legibilidade de textos em Língua Portuguesa (Version 3). arXiv. 10.48550/ARXIV.2203.12135

Okuhara, T., Furukawa, E., Okada, H., Yokota, R., & Kiuchi, T. (2025). Readability of written information for patients across 30 years: A systematic review of systematic reviews. Patient Education and Counseling, 135, 108656. 10.1016/j.pec.2025.108656

Orji, A., Kamenov, K., Dirac, M., Davis, A., Chadha, S., & Vos, T. (2020). Global and regional needs, unmet needs and access to hearing aids. International Journal of Audiology, 59(3), 166 –172. 10.1080/14992027.2020.1721577

Paaß, G., & Giesselbach, S. (2023). Foundation Models for Natural Language Processing: Pre-trained Language Models Integrating Media. Springer International Publishing. 10.1007/978-3-031-23190-2

Park, Y.-J., Pillai, A., Deng, J., Guo, E., Gupta, M., Paget, M., & Naugler, C. (2024). Assessing the research landscape and clinical utility of large language models: A scoping review. BMC Medical Informatics and Decision Making, 24(1), 72. 10.1186/s12911-024-02459-6

Peng, C., Yang, X., Chen, A., Smith, K. E., PourNejatian, N., Costa, A. B., Martin, C., Flores, M. G., Zhang, Y., Magoc, T., Lipori, G., Mitchell, D. A., Ospina, N. S., Ahmed, M. M., Hogan, W. R., Shenkman, E. A., Guo, Y., Bian, J., & Wu, Y. (2023). A study of generative large language model for medical research and healthcare. Npj Digital Medicine, 6(1), 210. 10.1038/s41746-023-00958-w

Ratanjee-Vanmali, H., Swanepoel, D. W., & Laplante-Lévesque, A. (2019). Characteristics, behaviours and readiness of persons seeking hearing healthcare online. International Journal of Audiology, 58(2), 107–115. 10.1080/14992027.2018.1516895

Sallam, M. (2023). ChatGPT Utility in Healthcare Education, Research, and Practice: Systematic Review on the Promising Perspectives and Valid Concerns. Healthcare, 11(6), 887. 10.3390/healthcare11060887

Seitz, L., Bekmeier-Feuerhahn, S., & Gohil, K. (2022). Can we trust a chatbot like a physician? A qualitative study on understanding the emergence of trust toward diagnostic chatbots. International Journal of Human-Computer Studies, 165, 102848. 10.1016/j.ijhcs.2022.102848

Shi, Y., Zi, X., Shi, Z., Zhang, H., Wu, Q., & Xu, M. (2024). *Enhancing Retrieval and Managing Retrieval: A Four-Module Synergy for Improved Quality and Efficiency in RAG System*s (Version 1). arXiv. 10.48550/ARXIV.2407.10670

Shool, S., Adimi, S., Saboori Amleshi, R., Bitaraf, E., Golpira, R., & Tara, M. (2025). A systematic review of large language model (LLM) evaluations in clinical medicine. BMC Medical Informatics and Decision Making, 25(1), 117. 10.1186/s12911-025-02954-4

Shumailov, I., Shumaylov, Z., Zhao, Y., Papernot, N., Anderson, R., & Gal, Y. (2024). AI models collapse when trained on recursively generated data. Nature, 631(8022), 755–759. 10.1038/s41586-024-07566-y

Singh, S., Jamal, A., & Qureshi, F. (2024). Readability Metrics in Patient Education: Where Do We Innovate? Clinics and Practice, 14(6), 2341–2349. 10.3390/clinpract14060183

Singhal, K., Azizi, S., Tu, T., Mahdavi, S. S., Wei, J., Chung, H. W., Scales, N., Tanwani, A., Cole-Lewis, H., Pfohl, S., Payne, P., Seneviratne, M., Gamble, P., Kelly, C., Babiker, A., Schärli, N., Chowdhery, A., Mansfield, P., Demner-Fushman, D.,…Natarajan, V. (2023). Large language models encode clinical knowledge. Nature, 620(7972), 172–180. 10.1038/s41586-023-06291-2

Siriwardhana, S., Weerasekera, R., Wen, E., Kaluarachchi, T., Rana, R., & Nanayakkara, S. (2023). Improving the Domain Adaptation of Retrieval Augmented Generation (RAG) Models for Open Domain Question Answering. Transactions of the Association for Computational Linguistics, 11, 1–17. 10.1162/tacl_a_00530

Soong, D., Sridhar, S., Si, H., Wagner, J.-S., Sá, A. C. C., Yu, C. Y., Karagoz, K., Guan, M., Kumar, S., Hamadeh, H., & Higgs, B. W. (2024). Improving accuracy of GPT-3/4 results on biomedical data using a retrieval-augmented language model. PLOS Digital Health, 3(8), e0000568. 10.1371/journal.pdig.0000568

Stojanov, A. (2023). Learning with ChatGPT 3.5 as a more knowledgeable other: An autoethnographic study. International Journal of Educational Technology in Higher Education, 20(1), 35. 10.1186/s41239-023-00404-7

Swanepoel, D. W., & Manchaiah, V. (2024). The Transformative Potential of AI Chatbots in Hearing Health Care. The Hearing Journal, 77(7), 1,4. 10.1097/01.HJ.0001025988.40308.e1

Swanepoel, D. W., Manchaiah, V., & Wasmann, J.-W. A. (2023). The Rise of AI Chatbots in Hearing Health Care. The Hearing Journal, 76(04), 26,30,32. 10.1097/01.HJ.0000927336.03567.3e

The World Bank, UNESCO, UNICEF, USAID, FSDO, & Bill and Melinda Gates Foundation. (2022). The State of Global Learning Poverty: 2022 Update. Population and Development Review, 48(4), 1215–1216. 10.1111/padr.12534

Thirunavukarasu, A. J., Mahmood, S., Malem, A., Foster, W. P., Sanghera, R., Hassan, R., Zhou, S., Wong, S. W., Wong, Y. L., Chong, Y. J., Shakeel, A., Chang, Y.-H., Tan, B. K. J., Jain, N., Tan, T. F., Rauz, S., Ting, D. S. W., & Ting, D. S. J. (2024). Large language models approach expert-level clinical knowledge and reasoning in ophthalmology: A head-to-head cross-sectional study. PLOS Digital Health, 3(4), e0000341. 10.1371/journal.pdig.0000341

Tran, E. D., Vaisbuch, Y., Qian, Z. J., Fitzgerald, M. B., & Megwalu, U. C. (2021). Health Literacy and Hearing Healthcare Use. The Laryngoscope, 131(5). 10.1002/lary.29313

Tsimpida, D., Rajasingam, S., Panagioti, M., & Henshaw, H. (2024). The leaky pipeline of hearing care: Primary to secondary care evidence from the English Longitudinal Study of Ageing (ELSA). International Journal of Audiology, 63(5), 349–357. 10.1080/14992027.2023.2186814

Vaswani, A., Shazeer, N., Parmar, N., Uszkoreit, J., Jones, L., Gomez, A. N., Kaiser, L., & Polosukhin, I. (2017). *Attention Is All You Need* (Version 7). arXiv. 10.48550/ARXIV.1706.03762

Vetter, M. A., Jiang, J., & McDowell, Z. J. (2025). An endangered species: How LLMs threaten Wikipedia’s sustainability. AI & SOCIETY, 40(6), 4309–4322. 10.1007/s00146-025-02199-9

Wasmann, J.-W., Pragt, L., Eikelboom, R., & Swanepoel, D. W. (2022). Digital Approaches to Automated and Machine Learning Assessments of Hearing: Scoping Review. Journal of Medical Internet Research, 24(2), e32581. 10.2196/32581

Wikimedia Foundation. (2025, October 29). Wikipedia Recognized as a Digital Public Good. *Wikimedia Foundation*. https://wikimediafoundation.org/news/2025/10/29/wikipedia-recognized-as-a-digital-public-good/

Wilson, B. S., Tucci, D. L., Merson, M. H., & O’Donoghue, G. M. (2017). Global hearing health care: New findings and perspectives. The Lancet, 390(10111), 2503–2515. 10.1016/S0140-6736(17)31073-5

Wilson, B. S., Tucci, D. L., Moses, D. A., Chang, E. F., Young, N. M., Zeng, F.-G., Lesica, N. A., Bur, A. M., Kavookjian, H., Mussatto, C., Penn, J., Goodwin, S., Kraft, S., Wang, G., Cohen, J. M., Ginsburg, G. S., Dawson, G., & Francis, H. W. (2022). Harnessing the Power of Artificial Intelligence in Otolaryngology and the Communication Sciences. Journal of the Association for Research in Otolaryngology, 23(3), 319–349. 10.1007/s10162-022-00846-2

World Health Organization. (2020). The World Health Organization and Wikimedia Foundation expand access to trusted information about COVID-19 on Wikipedia. World Health Organization. https://www.who.int/news/item/22-10-2020-the-world-health-organization-and-wikimedia-foundation-expand-access-to-trusted-information-about-covid-19-on-wikipedia

World Health Organization. (2021). *World Report on Hearing* (1st ed). World Health Organization.

World Health Organization. (2024). Hearing aid service delivery approaches for low-and middle-income settings. https://www.who.int/publications/i/item/9789240087927

Wright, D., Masud, S., Moore, J., Yadav, S., Antoniak, M., Christensen, P. E., Park, C. Y., & Augenstein, I. (2025). *Epistemic Diversity and Knowledge Collapse in Large Language Models* (Version 6). arXiv. 10.48550/ARXIV.2510.04226

Yan, M., Cerri, G. G., & Moraes, F. Y. (2023). ChatGPT and medicine: How AI language models are shaping the future and health related careers. Nature Biotechnology, 41(11), 1657–1658. 10.1038/s41587-023-02011-3

Younis, H. A., Eisa, T. A. E., Nasser, M., Sahib, T. M., Noor, A. A., Alyasiri, O. M., Salisu, S., Hayder, I. M., & Younis, H. A. (2024). A Systematic Review and Meta-Analysis of Artificial Intelligence Tools in Medicine and Healthcare: Applications, Considerations, Limitations, Motivation and Challenges. Diagnostics, 14(1), 109. 10.3390/diagnostics14010109

Zhao, W. X., Zhou, K., Li, J., Tang, T., Wang, X., Hou, Y., Min, Y., Zhang, B., Zhang, J., Dong, Z., Du, Y., Yang, C., Chen, Y., Chen, Z., Jiang, J., Ren, R., Li, Y., Tang, X., Liu, Z.,…Wen, J.-R. (2023). *A Survey of Large Language Models* (Version 16). arXiv. 10.48550/ARXIV.2303.18223

